# Dengue vaccine acceptability in Peru: A mixed-methods study in two dengue-endemic Peruvian cities

**DOI:** 10.1101/2025.09.17.25335974

**Authors:** Emma B. Ortega, Jorge L. Cañari-Casaño, Alfonso S. Vizcarra, Roberto Camizan-Castro, E. Jennifer Ríos, Jhonny J. Córdova López López, Cristina Hidalgo, Luz M. Moyano, Amy C. Morrison, Valerie A. Paz-Soldan

## Abstract

**Introduction:** Dengue poses a major public health challenge in Peru, with Piura and Loreto experiencing recurrent outbreaks and limited control options. A dengue vaccine could complement current vector control strategies and reduce transmission, yet community perceptions and potential barriers to uptake remain unclear.

**Methodology/Principal Findings:** Our mixed-methods study design was conducted in the dengue-endemic districts of Piura and Iquitos, and was guided by the 5C model (confidence, complacency, convenience, communication, and context). We conducted sixteen focus group discussions (147 participants) to explore perceptions, motivations, and concerns regarding a hypothetical future dengue vaccine. Next, a survey (n=883) assessed willingness to receive the vaccine, and the factors associated with vaccine refusal, using logistic regression.

**Results:** Overall, 81.2% of survey participants indicated willingness to receive a dengue vaccine. Qualitative findings underscored the need for clear information on efficacy, eligibility, and side effects, provided by trusted health professionals. Survey participants wanted to know about vaccine side effects (83.1%), effectiveness (69.8%), and number of doses (33.8%) to feel confident about the vaccine. In multivariate analysis, lack of knowledge about dengue transmission (OR 3.34), a negative opinion on the COVID-19 vaccine (OR 2.52), higher education levels (OR 6.48), and preference for natural immunity from an infection (OR 2.46) were associated with an increased risk of hesitancy towards the dengue vaccine. Willingness to pay for the vaccine (OR 0.37) and preference for house-by-house vaccination campaigns (OR 0.54) were associated with reduced levels of hesitancy.

**Conclusions:** The most important factors surrounding vaccine hesitancy were contained within confidence and context. While most participants were receptive to a future dengue vaccine, misinformation, negative COVID-19 vaccine views, and distrust of public institutions pose barriers. Tailored communication, engaging trusted local leaders, and ensuring easy access will be critical for successful dengue vaccination campaigns in these endemic regions.

**Author Summary:** Dengue, a mosquito-borne disease, is a growing global problem evidenced by increased outbreaks in recent years. Peru is a dengue-endemic country, with areas such as Iquitos and Piura consistently reporting high numbers of dengue cases every year. The main strategy for dengue control consists of reducing mosquito populations, including fumigation to kill adults and larvicide to prevent larva from developing into adults. Several dengue vaccines candidates provide promising support for reducing the burden of dengue, but community attitudes towards a future vaccine need to be examined so that future implementation can be successful. Through focus group discussions and surveys, our results found that themes of confidence and context influenced perceptions of a vaccine the most. Future vaccine campaigns should ensure messaging is clear and consistent, with information about vaccine side effects, eligibility, and doses. Outreach strategies should be implemented with accessibility in mind, as this will prevent barriers of community access.

## INTRODUCTION

Dengue is the most rapidly advancing vector-borne disease globally, causing more human morbidity and mortality worldwide than any other arthropod-borne virus [1–5]. In 2023 and 2024 record dengue outbreak numbers were the worst ever reported, with 256,641 reported cases in 2023 and 271,531 in 2024 [6]. The outbreak affected endemic areas, such as the states of Loreto (northern Amazon rainforest) and Piura (north coast), which have had the highest burden historically, but also areas that previously did not report significant transmission, like the capital city of Lima [6].

Vector control has traditionally been the primary public health strategy for the prevention and control of dengue, focused on eliminating larval habitats through source reduction (i.e., container removal) and larviciding to reduce *Aedes aegypti* and *Ae. albopictus* vectors, as well as using emergency measures with chemical pesticides (fumigation) to kill adult mosquitoes and mitigate dengue virus (DENV) transmission [7,8]. These strategies are challenging and costly to implement effectively. A dengue vaccine could complement these efforts by increasing immunity of human populations in at-risk areas which increase entomological thresholds (number of mosquitoes needed to) *sustain Aedes*-borne virus (ABV) transmission.

For decades, vaccine development for dengue has faced technical and safety challenges [9,10]. Currently, three vaccine candidates are either licensed or in advanced phase III clinical trials [9]. Dengvaxia (CYD-TDV, Sanofi Pasteur), partly tested in Piura, Peru, during phase II [11] has recently stopped production due to a lack of demand [12]. Recently, Qdenga (TAK003), has been approved in 40 countries and is available in 27 [13], including Peru, despite the company’s withdrawal of its Biologics License Application (BLA) from the U.S. FDA [14]. The Peruvian government began roll-out of the Qdenga vaccine in four dengue endemic regions of Peru in November 2024, including the states of Loreto and Piura. A third vaccine, the Butantan DV, developed by the Brazilian Butantan Institute and U.S. National Institutes of Health (NIH) is next in line for licensure. Phase III trials for the Butantan-DV vaccine have been completed [15], but is not yet in use because it is under regulatory review and awaiting approval. Merck, Sharp & Dohme (MSD, or “Merck” in the US) has also initiated Phase III of a single-dose V181 formulation [9].

Vaccinations are among the greatest public health advancements, significantly reducing the global burden of infectious diseases and childhood illnesses[16]. Attitudes toward vaccines, however, vary along a spectrum—from acceptance to refusal—with a gradient of hesitancy in between. Vaccine hesitancy reflects uncertainty driven by risk perception, concerns about safety, and mistrust of scientific institutions, many of which are related to access and knowledge [17]. Misinformation during the COVID-19 pandemic, vaccine uptake decreased globally but also contributed to hesitancy and push-back towards other vaccines, particularly in the Global North [18]. Although vaccine hesitancy remains less pronounced in Peru and other dengue endemic countries, compared to the United States, as dengue vaccines are rolled out [17], a clear assessment of issues associated with vaccine hesitancy and acceptability at both the community and health systems level urgently needed.

Our study used a mixed-method approach to assess motivations and barriers for uptake of a future dengue vaccine and to propose strategies for dengue vaccine deployment in Peru. At the time our study was conducted no dengue vaccine was commercially available so COVID-19 vaccination efforts in Peru were used to provide a framework for understanding public perceptions. To analyze and structure our findings, we employed the *‘5 C’s’* model of vaccine hesitancy, initially proposed by the Strategic Advisory Group of Experts on Immunization (SAGE) [19], and later updated by Razai et al. [20]. This model identifies five key factors influencing vaccine hesitancy: confidence, complacency, convenience, communication, and context. We present findings from focus group discussions and surveys conducted in two dengue-endemic Peruvian communities, exploring their willingness to adopt a dengue vaccine if available.

## METHODS

### Study Design

Our mixed-methods study started with a qualitative focus group discussions (FGDs) to identify barriers to community acceptance of a dengue vaccine and motivators for uptake based on the ‘5 C’s’ model [20]. Informed by these discussions, we created and administered a quantitative survey to evaluate the acceptance of a new dengue vaccine and to determine the main factors associated with distinct levels of vaccine hesitancy.

### Setting

We worked in two regions in Peru characterized by high dengue transmission and severe outbreaks: state of Piura in Northwestern coastal Peru and Iquitos City located Northeastern Peruvian Amazon basin. Piura reported the highest number of dengue cases in the country over the past 15 years (111,889 cases from 2007 to 2022) and ranks fifth in cumulative incidence of dengue with warning signs and severe dengue (722 cases per 100,000 inhabitants) [21]. In 2017, the El Niño phenomenon led to increased temperatures, rainfall, and flooding, triggering a major outbreak that accounted for 65% of all reported cases nationwide that year (68,290 cases) [22].

In 2023 and 2024, Piura became the epicenter of the largest dengue outbreaks in Peru, with 79,304 and 33,612 cases, respectively. In contrast, Iquitos bears the highest dengue burden in the Loreto region [6], which ranks second nationally in cumulative incidence of dengue with warning signs and severe dengue (1518 cases per 100,000 inhabitants) from 2007 to 2022 [21]. In 2023 and 2024, dengue also spread to Lima and southern regions, historically considered low-incidence areas [6].

We started our study in Iquitos City, the capital of Loreto Department, with a population of approximately 400,000 residents is surrounded on three sides by the Itaya, Nanay, and Amazon rivers and is accessible only by boat or plane, forming a socio-epidemiological island [23]. The city is divided into four districts, each of which also have rural sections along the Iquitos-Nauta highway and adjacent rivers. Additionally, the city has been a dengue research hub since the early 1990s, Iquitos has been a major hub for dengue research, contributing significantly to scientific understanding in multiple areas, including the spatiotemporal dynamics, behavior, and genetic structure of the *Aedes aegypti* vector [24–32]; the community-level understanding and lived experience of dengue illness [33,34]; the evaluation of targeted and city-wide vector control strategies [35–39]; the longitudinal and interepidemic transmission patterns of dengue virus [40]; the feasibility of implementing community-based surveillance tools for febrile disease detection [41–43]; and the role of human mobility in shaping transmission risk and spatial spread [44–49]; among other relevant contributions to the field [50,51]. Based on our studies, dengue virus activity remains elevated from August to April, peaking between November and January [39,52]. Until late 2019, DENV-2 was the dominant serotype; thereafter, DENV-1 emerged, triggering an outbreak that was overshadowed by the COVID-19 pandemic (http://dx.doi.org/10.4269/ajtmh.22-0539). According to the Peruvian National Institute of Statistics (INEI), 30% of the urban population in the Amazon rainforest has at least one unmet basic need, and 18.2% lives in poverty [53]. Despite these structural challenges, literacy rates remain relatively high, ranging from 88% to 92% [23]. Our study was carried out in the Punchana and Iquitos districts of Iquitos City.

We then moved to Piura in the northern tropical coastal region of Peru, which has shown a markedly different pattern of dengue virus behavior compared to Iquitos. With Piura Department, with an estimated population of 1,856809 residents [23], we worked in Chulucanas and La Matanza districts (Morropón Province) and La Unión district (Piura Province).

### Data Collection Instruments

To facilitate discussion in the FGDs, we used COVID-19 vaccination as a starting point, as participants had all recently been engaged in vaccination programs. Participants were then asked to envision a scenario where the Ministry of Health had approved a dengue vaccine and addressed three key questions: 1) What information would you need to accept vaccination? 2) How can public confidence in the vaccine be improved? 3) What is the best way to distribute a new vaccine quickly? The Iquitos study team developed and piloted the FGD guides in Spanish.

A survey was then developed using the preliminary results from the FGDs and the Vaccination Attitudes Examination (VAX) scale [54]. This 12-item VAX scale, validated in Spanish, measured four constructs: “1) mistrust of vaccine benefit, 2) worries about unforeseen future effects, 3) concerns about commercial profiteering, and 4) preference for natural immunity” [54]. After piloting this instrument with 20 individuals, our final revision included adaptation of the VAX scale from a 7-point to a 5-point Likert scale for ease of use. The final survey included topics covering: 1) participant sociodemographic information; 2) past experiences with COVID-19 and dengue; 3) COVID-19 vaccine use and perceptions; 4) VAX scale; 5) questions that addressed the 5C’s model; 6) participation in vector control activities; and 7) knowledge, attitudes, and practices regarding dengue and its prevention. The survey was developed, collected, and managed using REDCap (version 14.0.8) electronic data capture tools [55,56] and applied by our team on tablets.

### Recruitment and Procedures

Both qualitative (FGDs) and quantitative (survey) components of our study was stratified by intervention and control groups. In the city of Iquitos, the intervention group was defined as clusters (grouped blocks) that had previous participated in a vector control trial [36] whereas the control groups were selected from clusters where residents had no prior exposure to community-based research studies conducted by our group. Both the intervention and control groups were in Iquitos and Punchana districts of the city. We replicated this process in Piura, with the intervention group defined as districts where the Dengvaxia vaccine trial took place, located in the Chulucanas and La Matanza districts [11]. Each intervention group was matched to a control area in La Unión district, which was not involved in the vaccine trial.

We recruited FGD participants from a 5×5 block radius through door-to-door convenience sampling one day prior to the FGDs, limiting participation to two participants per block. Eligible participants were adults (>18 years old) who made household decisions, were mentally competent to provide consent, and met the age and gender criteria for the selected FGD. The research team returned 1–2 hours before each FGD to assist with transportation to the site. The FGD team was led by a senior qualitative researcher (VPS) and three research assistants: one co-facilitated (ASVS), one recorded participant feedback on butcher paper to facilitate discussion (EJRL in Iquitos, CC in Piura), and the third took notes (JLCC). We conducted 16 focus group discussions (FGDs) between February and March 2022—eight in Iquitos, followed by eight in Piura. In both study sites, four FGDs were conducted with residents from intervention areas and four with residents from control areas [36]. We stratified all FGDs by gender (female or male) and age (20–39 or 40–60 years). All FGDs were audio-recorded.

To meet our quantitative survey goal of 400 participants per study area, we used random sampling to select blocks in both the intervention and control areas to target recruitment in every household within those blocks. Recruitment was door to door until target enrollment was reached of eligible adult participants (aged 20-60) who were household decision makers. ASVS oversaw training and field monitoring and once data collection started, field work; JLCC managed quality control as data was entered on tablets and uploaded at least twice a day. Before starting data collection, the interviewers were piloted and trained in survey administration and research ethics. The surveys were conducted in August in Iquitos and in September 2022 in Piura, targeting heads of household between 20 and 60 years of age.

### Data Analysis

Audio recordings of our FGDs were transcribed into Spanish using the Trint platform [57]. Subsequently our research team conducted quality control (editing, verification, de-identification) of the transcriptions within the Trint software [57]. The 5 C’s ––confidence, complacency, convenience, communication, and context – were used as deductive codes, and emerging themes were added as subcodes inductively. Data from the FGDs was managed, coded, and analyzed using Dedoose Version 9.0.107 software [58].

For our quantitative survey, our main outcome of interest –– “acceptability of a new dengue vaccine” – was based on the following context where all questions were framed as *“If a dengue vaccine were available, what would you do?”* Responses adapted from the Oxford COVID-19 vaccine hesitancy scale [59], included: a) I would get it as soon as I can, b) I would get it when I have time, c) I would delay getting it, d) I would avoid getting it, and e) I would never get it. For this analysis, the variable was dichotomized into ‘willing to receive’ (responses a and b) and ‘not willing to receive’ (responses c, d, and e).

We conducted a descriptive analysis of categorical variables and a bivariate analysis between our main outcome of interest and other variables of interest using proportions and the chi-square test. To estimate socioeconomic status (SES), we applied principal component analysis based on household goods and services [60]. Each participant received an SES score, categorized into low, medium, and high tertiles. To identify factors associated with non-acceptance (not willing to receive) of a new dengue vaccine, logistic regression and estimated adjusted odds ratios (OR) were used. The model adjusted for sociodemographic factors, dengue knowledge and prevention practices, experiences with dengue and severe COVID-19-as they related to the *‘5 C’s’* model of vaccine hesitancy, and variables from the vaccination attitudes scale. Data was cleaned using Stata 15.0 (Stata Corp., College Station, Texas, United States), whereas analysis and graphics were created in R version (Version 4.4.3; R Foundation for Statistical Computing).

### Ethics Statement

Our study protocol was approved by the Institutional Review Boards (IRBs) of PRISMA (CE0882.21) providing local Peruvian approval and the Tulane School of Public Health and Tropical Medicine (2021-1749). The University of California, Davis IRB provided an exemption. The protocol was also reviewed and approved by the Loreto Regional Health Department which oversees health research Iquitos. Written consent was obtained from all participants prior to the initiation of FGDs and to surveys.

## RESULTS

### Study Population

A total of 147 individuals participated in the 16 FGDs: 72 from Iquitos and 75 from Piura. Over half of participants were female (53%), and 52% were aged 20-39. In 15 of the 16 FGDs, at least one participant had experienced dengue. Most participants had received at least one dose of a COVID-19 vaccine [Table 1]. The full table of quotes in English and the original Spanish can be found in the supplemental files [S1 Table].

**Table 1.** Focus Group Discussion Population Characteristics.

| Site | Gender | Age Group | Number of Participants | Participants who have had dengue | Participants with at least ≥1 COVID-19 Vaccine Dose |
| --- | --- | --- | --- | --- | --- |
| Iquitos<br>(n=72)<br>(8 GFs) | Male | 20–39 | 16 | 6 | >9 |
|  |  | 40–60 | 17 | 5 | 17 |
|  | Female | 20–39 | 19 | 1 | 18 |
|  |  | 40–60 | 20 | 11 | 19 |
| Piura<br>(n=75)<br>(8 GFs) | Male | 20–39 | 23 | 7 | 23 |
|  |  | 40–60 | 14 | 2 | 14 |
|  | Female | 20–39 | 19 | 7 | 19 |
|  |  | 40–60 | 19 | 2 | 19 |
| TOTAL |  |  | 147 | 44 | 138 |

*In Iquitos, data on COVID-19 vaccination was not recorded for one male focus group discussion aged 20–39 (represented as “>9”).*

The survey included 883 participants, with a similar site breakdown of 49% (432) from Iquitos and 51% (451) from Piura. The household refusal rate was higher in Piura (29%, 186 of 637 approached did not participate), compared to 23% (132 of 564) in Iquitos; the primary reasons given for not participating was time constraints and not wanting to signature (on the consent form). Nearly half of participants were aged 40-60 (48.7%), the majority were women (71.1%), and around 80% had completed secondary education or higher [Table 2].

**Table 2.** Sociodemographic Characteristics of Survey Participants.

| Characteristic | Overall, N = 883<sup>1</sup> | Iquitos, N = 432<sup>1</sup> | Piura, N = 451<sup>1</sup> | p-value<sup>2</sup> |
| --- | --- | --- | --- | --- |
| Age |  |  |  | 0.13 |
| 18 to 39 years old | 51.3% (453/883) | 53.9% (233/432) | 48.8% (220/451) |  |
| 40 to 60 years old | 48.7% (430/883) | 46.1% (199/432) | 51.2% (231/451) |  |
| Gender |  |  |  | <0.001 |
| Female | 71.1% (628/883) | 63.2% (273/432) | 78.7% (355/451) |  |
| Male | 28.9% (255/883) | 36.8% (159/432) | 21.3% (96/451) |  |
| Education Level |  |  |  | <0.001 |
| Primary school | 16.4% (145/883) | 3.5% (15/432) | 28.8% (130/451) |  |
| High school | 45.6% (403/883) | 46.5% (201/432) | 44.8% (202/451) |  |
| Technical and/or higher | 37.9% (335/883) | 50.0% (216/432) | 26.4% (119/451) |  |
| Wealth index terciles |  |  |  | <0.001 |
| 1 (poorer) | 33.5% (292/871) | 16.9% (72/426) | 49.4% (220/445) |  |
| 2 | 33.2% (289/871) | 31.2% (133/426) | 35.1% (156/445) |  |
| 3 (wealthier) | 33.3% (290/871) | 51.9% (221/426) | 15.5% (69/445) |  |
| Residence area |  |  |  | <0.001 |
| Urban | 81.7% (721/883) | 100.0% (432/432) | 64.1% (289/451) |  |
| Peri-urban | 18.3% (162/883) | 0.0% (0/432) | 35.9% (162/451) |  |
| Number of household members |  |  |  | <0.001 |
| 1 to 3 people | 22.3% (197/883) | 17.1% (74/432) | 27.3% (123/451) |  |
| 4 to 6 people | 53.8% (475/883) | 48.6% (210/432) | 58.8% (265/451) |  |
| 7 to more people | 23.9% (211/883) | 34.3% (148/432) | 14.0% (63/451) |  |
| Presence of child >5 years old | 40.4% (357/883) | 43.3% (187/432) | 37.7% (170/451) | 0.090 |
| Presence of older adults (Yes) | 39.0% (344/883) | 55.3% (239/432) | 23.3% (105/451) | <0.001 |
| Presence of vulnerable persons (pregnancy, chronic illness, physical disability) | 26.6% (235/883) | 34.5% (149/432) | 19.1% (86/451) | <0.001 |
| Close experience with severe dengue | 15.2% (134/883) | 19.9% (86/432) | 10.6% (48/451) | <0.001 |
| Willing to receive the dengue vaccine |  |  |  | <0.001 |
| Willing to receive | 81.2% (696/857) | 76.2% (327/429) | 86.2% (369/428) |  |
| In doubt or hesitancy | 18.8% (161/857) | 23.8% (102/429) | 13.8% (59/428) |  |

<sup>1</sup>% (n/N)
<sup>2</sup>Pearson's Chi-squared test
*5Cs Framework*

### 5Cs Framework

Below we present a summary and relevant quotes for findings, organized under the 5C’s model.

### Confidence

Both qualitative and quantitative data indicated generally positive attitudes towards a potential dengue vaccine, and the scales showed relatively low vaccine hesitancy [Tables 2 and 3]. The following statements from FGD reflected this sentiment: *“We assume that a vaccine is good for your health. Who would not want to get it? But always as long as we know what symptoms [side effects] we will get. Right?”* (FG1). Survey results were consistent with FGDs findings, with 81.2% (696) of respondents indicating they were willing to receive a new dengue vaccine when available. Two main themes emerged in FGDs regarding confidence about a new vaccine: 1) the need for clear information about the vaccine to increase people’s confidence in it, including information on its efficacy, number of doses required for protection, and possible side effects, and 2) vaccine information should come from trusted institutions, naming the Ministry of Health and health professionals as such [Table 3].

**Table 3.** Themes Identified in the FGDs and Relevant Quotes.

| Themes | Subcodes | Quotes |
| --- | --- | --- |
| **Section: Confidence** |  |  |
| Availability of clear and transparent information on the vaccine | Vaccine side effects | *We imagine that it's a vaccine which is good for your health. Who wouldn't want to get it? But as long as they end up knowing what symptoms we're going to have. Right? (FG1)*<br>*And the fear of a reaction to the vaccine. (FG1)* |
|  | Vaccine eligibility | *Have they tested it on a person who's had dengue, including a severe case? (FG9)*<br>*What problems would it cause in a pregnant woman, or a diabetic person, or for any other disease a person may have? (FG10)* |
|  | Evidence from others | *First, like I said, wait for others to get vaccinated, and see what reactions they have. (FG5)* |
|  | Number of doses | *Yes, because he already has three vaccines, three doses, a vaccine is never three doses, it's only two. (FG1)* |
|  | Science | *Because I always say that vaccines protect against the bad that's happening in these times. (FG1)*<br>*To protect ourselves from the disease and logically we have to get vaccinated, because if we are not vaccinated we are at risk for the disease (FG2)* |
|  | Immunity | *Depending on the guarantees they would give me- if it's immunization for life, then I would get it. (FG2)* |
| Experience with Dengue | Experience with Dengue | *Well, as someone from Loreto, I've had dengue at some point. (FG2)*<br>*I think the dengue vaccine is something that is highly anticipated and I think this event would be a boom. (FG2)*<br>*Because now it's different, because the diseases are evolving and they are no longer the same, I think. Now the disease is stronger, well that's what I think. (FG13)* |
| Trusted Healthcare Professionals | Local trusted sources | *Well, here in Unión, we honestly don't trust the authorities. What they promise are lies. Maybe they were telling me something that isn't true. (FG11)* |
|  | National sources | *The first thing I would do is find out if the Ministry of Health has approved it. (FG2)* |
| **Section: Complacency** |  |  |
| Reasons for Vaccinating | Self-perceived risk from past experience | *But when you get dengue hemorrhagic fever, you start to worry. That's when you start to realize it's not a joke, with dengue. (FG13)*<br>*Dengue has been around for many years, but what did we do? What did we use to cure dengue? Pure bitters, and we still use them. (FG13)* |
|  | Protecting the health of children | *It's a responsibility, as a parent, to get them vaccinated, so that they grow up healthy and strong with their childhood vaccines. (FG11)*<br>*Well, to me, they should vaccinate the little ones first, to avoid that dengue disease. (FG14)* |
|  | Work/ Travel and Policies | *Because to enroll in school they will force you to get the children vaccinated. (FG4)*<br>*To play football they are asking for the vaccine, if you don't have it, you can't play. (FG5)* |
| Reasons for Not Vaccinating | Low self-perceived risk | *It's never happened to me. I mean, I've never gotten sick enough to call a doctor. (FG6)*<br>*I'm not afraid because you just get a mild dengue infection, right? (FG13)* |
|  | Religious views | *For the simple reason that it says it is the sign of the beast. (FG1)* |
| Vector Control | Vaccine effect on vector control | *As for me, what I want to know is if I get vaccinated, then I no longer need to kill my mosquitoes? (FG2)*<br>*For dengue, we should put on repellent or put [a repellent device on wall] or use a mosquito coil, right? To protect ourselves, we should continue doing this, even if we've been vaccinated? (FG10)* |
| **Section: Convenience** |  |  |
| Physical Barriers | Long lines | *People got up early, brought their chairs, put them in front of the line, and they sold their spot for 10 soles or 20 soles. (FG1)* |
|  | Costs | *Generally, vaccines are provided by the government. I don't know of a vaccine that we can buy. (FG2)*<br>*I prefer to save my family, I can pay whatever they tell me to. 50, 20 soles, I'll pay it. It's to protect us, to save our lives. (FG11)* |
| Improving Access | Ideas for rolling out the dengue vaccine | *And sustained awareness is very important, not just going once and then not returning. To change the mental structure of the people in the community, it [awareness building] has to be constant, you have to be with them, try different [messages]. (FG3)* |
| **Section: Communication** |  |  |
| COVID-19 Misinformation | Misinformation | *If you're vaccinated you would become sterile or something like that, so she hasn't had a single vaccine (FG10)* |
| Vaccine Information | Sources of information | *I don't trust the vaccine for reasons I saw on the news. I let myself be influenced by the news (FG7)*<br>*The radio station, well, the station as well. (FG10)* |

To build confidence in the new vaccine, survey participants wanted information on potential adverse effects (83.1%), vaccine effectiveness (69.8%), the number of required doses (33.8%), and the manufacturing laboratory (9.1%) [Table 4]. Hesitant participants were particularly interested in information about where the vaccine was manufactured compared to their accepting counterparts (13.0% vs 8.2%, p = 0.054) [Table 4].

**Table 4.** Bivariate analysis of factors associated with *‘5 C’s’* model of vaccine hesitancy and acceptability of the new dengue vaccine.

| Characteristic | Overall,<br>N = 857<sup>1</sup> | Willing to<br>receive,<br>N = 696<sup>1</sup> | In doubt or<br>hesitant,<br>N = 161<sup>1</sup> | p-value<sup>2</sup> |
| --- | --- | --- | --- | --- |
| **Confidence** |  |  |  |  |
| *I would like to know:* |  |  |  |  |
| Its adverse effects | 83.1% (712/857) | 85.1% (592/696) | 74.5% (120/161) | 0.001 |
| How effective the vaccine is | 69.8% (598/857) | 70.5% (491/696) | 66.5% (107/161) | 0.3 |
| The number of doses required | 33.8% (290/857) | 35.2% (245/696) | 28.0% (45/161) | 0.080 |
| Which laboratory manufactured the vaccine | 9.1% (78/857) | 8.2% (57/696) | 13.0% (21/161) | 0.054 |
| **Complacency** |  |  |  |  |
| Permits fumigation in the home |  |  |  | <0.001 |
| Not reluctant | 79.5% (670/843) | 81.9% (562/686) | 68.8% (108/157) |  |
| Reluctant sometimes | 14.8% (125/843) | 13.6% (93/686) | 20.4% (32/157) |  |
| Always reluctant | 5.7% (48/843) | 4.5% (31/686) | 10.8% (17/157) |  |
| Permits focal treatment in the home |  |  |  | 0.13 |
| Not reluctant | 93.5% (799/855) | 94.3% (656/696) | 89.9% (143/159) |  |
| Reluctant sometimes | 4.0% (34/855) | 3.4% (24/696) | 6.3% (10/159) |  |
| Always reluctant | 2.6% (22/855) | 2.3% (16/696) | 3.8% (6/159) |  |
| Participates in campaigns to eliminate<br>larval habitat sites |  |  |  | 0.041 |
| Not reluctant | 45.9% (320/697) | 45.2% (252/557) | 48.6% (68/140) |  |
| Reluctant sometimes | 17.9% (125/697) | 19.7% (110/557) | 10.7% (15/140) |  |
| Always reluctant | 36.2% (252/697) | 35.0% (195/557) | 40.7% (57/140) |  |
| Close experience with severe dengue | 15.4% (132/857) | 15.2% (106/696) | 16.1% (26/161) | 0.8 |
| Close experience with severe COVID-19 | 36.3% (311/857) | 36.5% (254/696) | 35.4% (57/161) | 0.8 |
| COVID-19 vaccine doses received |  |  |  | <0.001 |
| Three or more doses | 84.1% (721/857) | 86.2% (600/696) | 75.2% (121/161) |  |
| None or incomplete (1-2) doses | 15.9% (136/857) | 13.8% (96/696) | 24.8% (40/161) |  |
| Opinion of the COVID-19 vaccine |  |  |  | <0.001 |
| In favor | 80.6% (691/857) | 85.5% (595/696) | 59.6% (96/161) |  |
| Against or in doubt | 19.4% (166/857) | 14.5% (101/696) | 40.4% (65/161) |  |
| **Convenience** |  |  |  |  |
| *I prefer they vaccinate by:* |  |  |  |  |
| House by house | 65.8% (564/857) | 67.4% (469/696) | 59.0% (95/161) | 0.043 |
| In schools | 20.1% (172/857) | 20.5% (143/696) | 18.0% (29/161) | 0.5 |
| At health centers or hospitals | 20.3% (174/857) | 21.0% (146/696) | 17.4% (28/161) | 0.3 |
| In public places | 17.7% (152/857) | 17.2% (120/696) | 19.9% (32/161) | 0.4 |
| I would be willing to pay for the vaccine | 41.2% (353/857) | 44.8% (312/696) | 25.5% (41/161) | <0.001 |
| **Context** |  |  |  |  |
| Previous experience in dengue research | 48.2% (413/857) | 49.1% (342/696) | 44.1% (71/161) | 0.2 |
| Study Site |  |  |  | <0.001 |
| Iquitos | 50.1% (429/857) | 47.0% (327/696) | 63.4% (102/161) |  |
| Piura | 49.9% (428/857) | 53.0% (369/696) | 36.6% (59/161) |  |
| Age |  |  |  | 0.10 |
| 18 to 39 years old | 51.2% (439/857) | 49.9% (347/696) | 57.1% (92/161) |  |
| 40 to 60 years old | 48.8% (418/857) | 50.1% (349/696) | 42.9% (69/161) |  |
| Gender |  |  |  | 0.041 |
| Female | 70.6% (605/857) | 72.1% (502/696) | 64.0% (103/161) |  |
| Male | 29.4% (252/857) | 27.9% (194/696) | 36.0% (58/161) |  |
| Education Level |  |  |  | <0.001 |
| Primary school | 16.1% (138/857) | 18.5% (129/696) | 5.6% (9/161) |  |
| High school | 45.2% (387/857) | 46.4% (323/696) | 39.8% (64/161) |  |
| Technical and/or higher | 38.7% (332/857) | 35.1% (244/696) | 54.7% (88/161) |  |
| Socioeconomic Level |  |  |  | 0.021 |
| 1 (poorer) | 33.1% (280/845) | 33.7% (231/686) | 30.8% (49/159) |  |
| 2 | 33.1% (280/845) | 34.7% (238/686) | 26.4% (42/159) |  |
| 3 (wealthier) | 33.7% (285/845) | 31.6% (217/686) | 42.8% (68/159) |  |
| Residence area |  |  |  | 0.002 |
| Urban | 82.1% (704/857) | 80.2% (558/696) | 90.7% (146/161) |  |
| Peri-urban | 17.9% (153/857) | 19.8% (138/696) | 9.3% (15/161) |  |
| *Household Composition* |  |  |  |  |
| Presence of child >5 years old (Yes) | 40.1% (344/857) | 40.9% (285/696) | 36.6% (59/161) | 0.3 |
| Presence of older adults (Yes) | 39.9% (342/857) | 39.2% (273/696) | 42.9% (69/161) | 0.4 |
| Presence of vulnerable persons<br>(pregnancy, chronic illness, physical<br>disability) | 27.0% (231/857) | 27.0% (188/696) | 26.7% (43/161) | >0.9 |

<sup>1</sup>% (n/N)
<sup>2</sup>Pearson's Chi-squared test; Fisher's exact test

Similar issues and questions revealing hesitancy among some participants emerged in the FGDs: these focused on participants wanting information on the vaccine’s components, duration of protection, potential side effects, and whether people in other areas or countries had already been vaccinated – or if they would be first: *“I will wait for others to get it, but I am not going to risk getting it first”* (FG11). There were questions regarding vaccine eligibility, ranging from having had dengue before, to being pregnant or having co-morbidities: *“What problems would it cause in a pregnant woman or a diabetic [person] or any other disease a person may have?”* (FG10).

In the vaccination attitudes examination scale (VAX), the top three statements that participants agreed with focused on trusting vaccines against severe infectious diseases (83%), feeling protected after being vaccinated (81%), and feeling safe after vaccinations (78%) [Figure 1]. Our multivariate model showed that participants who preferred natural immunity from an infection over a vaccine were less likely to want the dengue vaccine (OR 1.25, 95% CI 0.61 to 2.54) [table 5].

**Figure 1.**
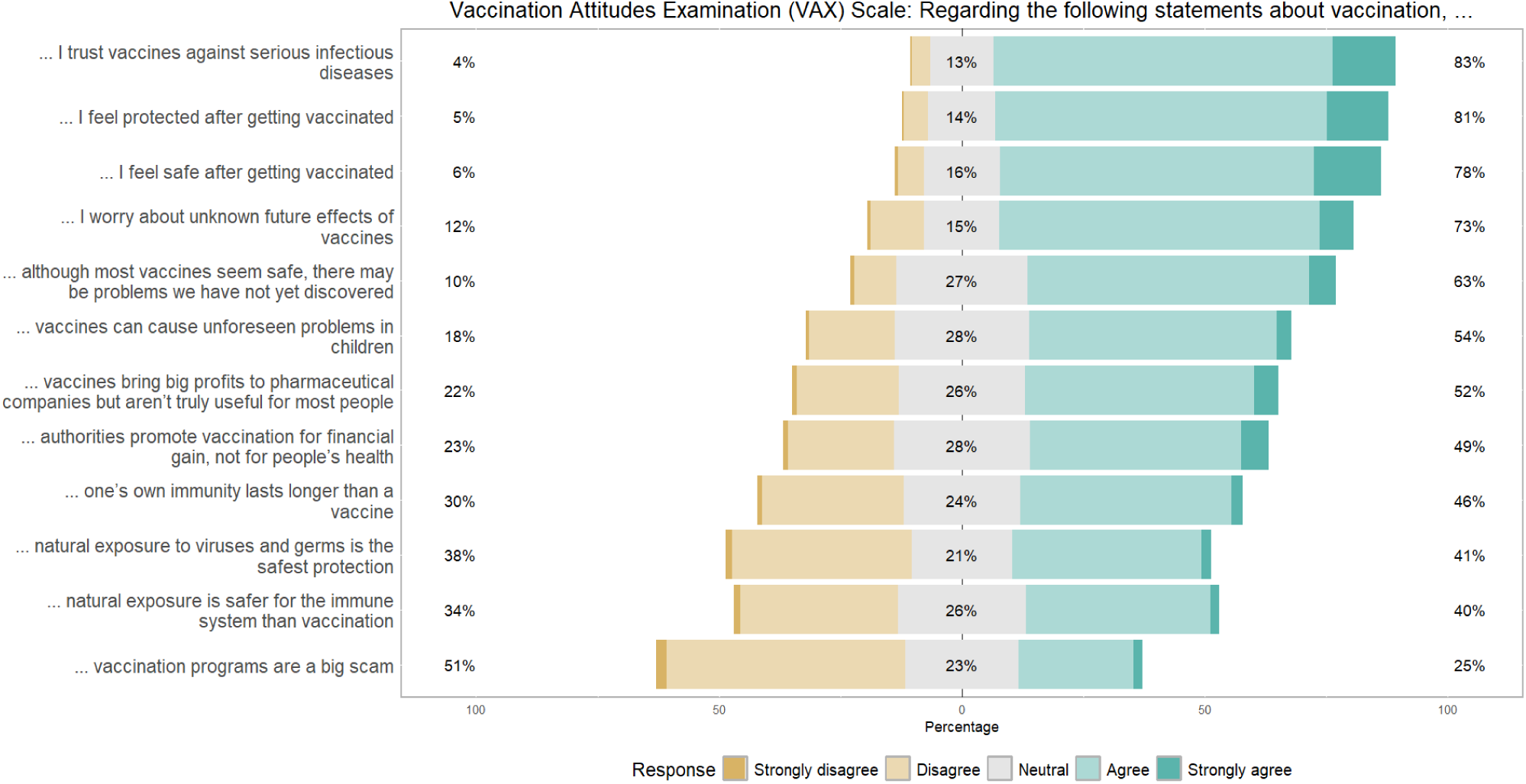
Vaccination Attitudes Examination (VAX) Scale.

**Table 5.** Multivariate Analysis Examining Factor Associated with Non-Acceptance (not willing to receive) of a new Dengue Vaccine.

| Characteristic | Not willing to receive the new dengue vaccine |  |  |
| --- | --- | --- | --- |
|  | OR<sup>1</sup> | 95% CI<sup>1</sup> | p-value |
| **'5 C's' model: Confidence** |  |  |  |
| Opinion of the COVID-19 vaccine |  |  | 0.001 |
| In favor | Ref. | — |  |
| Against or in doubt | 2.52 | 1.45, 4.42 |  |
| Know how dengue is transmitted (Yes) |  |  | 0.001 |
| Yes | Ref. | — |  |
| No | 3.34 | 1.61, 6.89 |  |
| **'5 C's' model: Convenience** |  |  |  |
| I preferred that they vaccinate house by house (Yes) |  |  | 0.030 |
| No | Ref. | — |  |
| Yes | 0.54 | 0.31, 0.94 |  |
| I would be willing to pay for the vaccine (Yes) |  |  | <0.001 |
| No | Ref. | — |  |
| Yes | 0.37 | 0.22, 0.63 |  |
| **'5 C's' model: Context** |  |  |  |
| Education Level |  |  | 0.001 |
| Primary school | Ref. | — |  |
| High school | 3.19 | 1.21, 9.66 |  |
| Technical and/or higher | 6.48 | 2.24, 21.4 |  |
| **VAX Scale** |  |  |  |
| Preference for natural immunity |  |  | 0.004 |
| Low | Ref. | — |  |
| Intermediate | 2.46 | 1.40, 4.43 |  |
| High | 1.25 | 0.61, 2.54 |  |
| No. Obs. | 609 |  |  |

<sup>1</sup>OR = Odds Ratio, CI = Confidence Interval

| Characteristic | Not willing to receive the new dengue vaccine |  |  |
| --- | --- | --- | --- |
|  | OR<sup>1</sup> | 95% CI<sup>1</sup> | p-value |

Figure 1 Caption: The 12 statements used to create the Vaccination Attitudes Examination (VAX) Scale along with participant responses.

### Complacency

Complacency, referring to a low perceived risk for the vaccine-preventable disease, was gauged based on people’s perceived risk for and severity of dengue. In our study sites, all FGD and survey participants had either had dengue or knew someone who had it. Additionally, 15.4% of survey participants reported experiencing or knowing someone with severe dengue [Table 4]. The perceived severity of dengue seemed to influence FGD participants’ attitudes toward vaccination, with some expressing a strong desire for protection and high interest in a future dengue vaccine, particularly to prevent severe disease: *“When you get severe dengue, then you start to worry, then you start to become more conscious that dengue is no joke!”* (FG 13). There was also recognition that one could get dengue multiple times, worthy of prevention: *“I would get the vaccine, because… there are four types of dengue… I would fear hemorrhagic dengue the most. Besides that, dengue comes every year or when the river grows in our region. I would get it [vaccine]”* (FG3). Mothers in the FGDs showed interest particularly in vaccinating their children, with one suggesting that vaccine eligibility should start with children. Even parents hesitant to vaccinate themselves acknowledged the importance of protecting their children as part of their parental duty.

There was also a significant association between participation in vector control campaigns and vaccine hesitancy: study participants who were willing to permit fumigation in their homes and participate in campaigns to eliminate larval development sites for dengue prevention were more likely to be accepting of the dengue vaccine [Table 4]. Not knowing how dengue is transmitted (via the bite of an infected mosquito) was found to be associated with an increase in hesitancy towards the dengue vaccine (OR 3.34, 95% CI 1.61 to 6.89) [Table 5]. In the FGD’s, people asked whether getting vaccinated would mean they would no longer need to participate in vector control activities: “*As for me, what I want to know is if I get vaccinated, then I no longer need to kill my mosquitoes?”* (FG2).

To explore potential hesitancy toward a hypothetical dengue vaccine, we asked participants to reflect on their decision-making process regarding a recent ‘new vaccine’—the COVID-19 vaccine—as a proxy for evaluating acceptance of novel immunization strategies. The reasons participants provided for receiving the COVID-19 vaccine—with 84.1% of survey respondents reporting 3 or more COVID-19 vaccine doses [Table 4] – may also offer insights into their potential reactions to a dengue vaccine. Reasons given for the COVID-19 vaccine in the FGDs included having high-risk household members, community pressure to do so, and feeling sufficiently informed to decide. Some FGD participants cited obligation due to Peruvian laws requiring proof of vaccination for travel, banking, grocery shopping, or work as their main reason for getting vaccinated – with some saying it was the nudge they needed to get it. Disease severity was explored in the surveys: 36.3% of participants had experienced or knew someone with severe COVID-19 [Table 4]. People who lacked confidence in the COVID-19 vaccine were more likely to express hesitancy towards a dengue vaccine (OR 2.52, 95% CI 1.45 to 4.42) [Table 5].

### Convenience

Participants in FGDs reported long waiting lines as a barrier to accessing the COVID-19 vaccine; some specified queuing for hours in the morning, which was particularly challenging for individuals with work or caregiving responsibilities. Some admitted to a “known practice” of hiring others to hold their place in line, though this option was not affordable for everyone: *“Yes, some people got up very early, took their chairs and sat at the front of the line, and then would sell their spot for 10-20 soles (US$3-6)”* (FG1). However, none stated long waiting lines as a reason not to get the vaccine.

Most survey respondents also believed the vaccine should be free, as with the COVID-19 vaccine, with less than half (41.2%) of those surveyed reporting willingness to pay for it [Table 4]. This willingness to pay was found to be significantly associated with a decreased risk of vaccine hesitancy in our model (OR 0.37, 95% CI 0.22 to 0.63) [table 5]. In the FGDs, it was pointed out that the vaccine should be covered by fees paid in taxes or part of government healthcare coverage. These participants also suggested prices ranging from ∼$1.40-$13.60 USD, if necessary. Some argued that the vaccine cost was justified compared to the expenses of treating severe or hemorrhagic dengue. One individual noted that even the cost of over-the-counter drugs for mild dengue could exceed the vaccine cost: *“because one might not only be in a hospital… Sometimes you will need to buy something [medication] for the headaches. For prevention, I might as well get a vaccine that costs 30 soles [∼US$ 8] and then I won’t have other expenses”* (FG1). Participants in FGDs also highlighted that large families would struggle to afford multiple doses and proposed sliding scale costs or discounts, such as ∼$8.15 USD per person or ∼$13.60 USD for two people.

To improve access to a future dengue vaccine, survey participants suggested strategies such as house-to-house vaccination (65.8%), vaccination at schools (20.1%), health posts or hospitals (20.3%), and public places (17.7%) [Table 4]. Specifically, our model showed people who preferred house-to-house vaccination were less likely to express hesitancy towards a dengue vaccine (OR 0.54, 95% CI 0.31-0.94) [table 5].

### Context

Previous participation in dengue research did not have an impact on survey participants’ vaccine hesitancy, but when comparing the two sites there was a higher level of hesitancy in Iquitos (63.4%) compared to Piura (36.6%) [Table 4]. Men were more likely to report that they were not willing to receive the dengue vaccine. There was also a higher level of dengue vaccine hesitancy among survey participants with a technical or higher education level, a wealthier socioeconomic status, and those living in an urban area. Specifically, participants with a technical or higher education level were more likely to express hesitancy towards a dengue vaccine compared to those with a primary level education (OR 6.48, 95% CI 2.24 to 21.4) [Table 5]. No differences in hesitancy were found were found between age groups, houses with children older than 5 years old, elderly residents, or among households with a member with a vulnerable condition, including pregnancy, chronic illness, or a physical disability.

Drawing on their experience with COVID-19 vaccination, FGD participants recommended establishing vaccination posts in central, frequented areas, such as city centers. Despite these suggestions, FGD and survey respondents also identified challenges in reaching certain populations, including individuals with specific religious affiliations (e.g., evangelical), indigenous communities; and, only mentioned in the survey, people living in peri-urban areas and of lower socioeconomic status [Table 6].

**Table 6.** Public Health Considerations for Vaccine Outreach.

| Variables | Total<br>N = 857<sup>1</sup> | Willing to<br>receive<br>N = 696<sup>1</sup> | Not Willing<br>to receive<br>N = 161<sup>1</sup> | p-value<sup>2</sup> |
| --- | --- | --- | --- | --- |
| ***It would be difficult to vaccinate:*** |  |  |  |  |
| People who live outside the city | 385 (44.9%) | 327 (47.0%) | 58 (36.0%) | 0.012 |
| Religious or evangelical people | 323 (37.7%) | 269 (38.6%) | 54 (33.5%) | 0.2 |
| Indigenous people | 234 (27.3%) | 187 (26.9%) | 47 (29.2%) | 0.6 |
| People with low economic status | 79 (9.2%) | 63 (9.1%) | 16 (9.9%) | 0.7 |
| ***Media platform to publicize the vaccine*** |  |  |  | 0.3 |
| Radio | 150 (17.8%) | 116 (16.9%) | 34 (21.8%) |  |
| TV | 119 (14.1%) | 95 (13.8%) | 24 (15.4%) |  |
| Social media | 225 (26.7%) | 185 (26.9%) | 40 (25.6%) |  |
| Loudspeaker | 261 (30.9%) | 222 (32.3%) | 39 (25.0%) |  |
| House-to-house notification | 89 (10.5%) | 70 (10.2%) | 19 (12.2%) |  |

<sup>1</sup>n (%)
<sup>2</sup>Pearson's Chi-squared test

### Communication

At both sites, FGD participants expressed trust in their physicians, healthcare friends, and health professionals or community health workers from local facilities to endorse a vaccine. They also stated they would be more likely to get vaccinated for dengue if the Ministry of Health approved it. In Iquitos, one name came up in every FGD: most valued the opinion of a local physician and parish priest who had been influential and demonstrated commitment to the community at the height of the COVID-19 pandemic. In contrast, Piura participants expressed distrust in their local leaders and felt abandoned by the local government. Of note, many FGD participants reported they had negative experiences with visiting government health facilities but wanted approval from these same organizations before receiving a future dengue vaccine.

Participants in FGDs noted that clear messaging is crucial, as changing COVID-19 vaccine recommendations had eroded their trust. Participants in FGDs described receiving and believing in information about COVID-19 vaccines from non-healthcare sources such as radio, television (local and national programs), news channels, internet sources (including social media), religious organizations, and family and friends. Relatedly, in most FGD, participants mentioned that health promotion messages must be tailored since sub-populations may face different barriers, specifying certain religious groups that had refused COVID-19 vaccination and indigenous populations that may need different outreach strategies.

To promote a dengue vaccine, FGD and survey respondents recommended used the radio (including regional programs such as *La Voz de la Selva, Radio Loreto, Radio Saby Star*, and national programs, such as *Radio Exitosa*), TV programs, social media like Facebook, and using a truck with speakers to drive around neighborhoods to share information (more commonly suggested in Piura where this method is used in some communities) [Table 6]. They also suggested press releases. For healthcare engagement, participants suggested holding open forum townhalls where health professionals could educate and answer questions.

## DISCUSSION

As dengue vaccines reach communities around the world, assessing the factors that may impede their acceptability and uptake in high-risk communities prior to dissemination is key. Vaccine acceptance was high in these two dengue endemic regions of Peru, and most FGD and survey participants revealed positive attitudes towards a potential dengue vaccine. Our study population had a baseline dengue vaccine acceptance of about 81.2%, consistent with reports in other countries [61,62]. We used the five Cs model to examine which categories were most associated with willingness to participate in vaccination programs, and it was found that confidence and context factors were most associated with hesitancy.

Based on the importance of confidence, communication needs to be clear and effective. A potential policy implication for MINSA includes adding the vaccine to the national immunization schedule. This would allow clinicians to recommend the dengue vaccine to their patients at routine visits, capitalizing on patients pre-existing trust in health professionals. The results highlight the importance of identifying who the community turns to for their health advice. These individuals can be chosen as program champions to promote messaging about a dengue vaccine. For example, Padre Raymundo earned the trust of community members at the beginning of the COVID-19 pandemic, when he raised money to buy oxygen for the local hospitals who had run out [63]. Due to this, he is well-known and trusted in Iquitos. Taking it a step further, if the program champions selected have received the dengue vaccine themselves, people may have greater acceptance to receive it due to seeing evidence of any side effects and not being the first ones vaccinated. Various methods of communication could be used to promote these campaigns, including radio, television, and social media, to ensure the message is spread to the entire community.

Within context, participants identified hard to reach groups, which may require special attention for vaccine roll-out. Lessons can be learned from the rollout of COVID-19: long lines, limited hours of availability, and transportation availability created physical barriers towards immunization. Communities outside of the city limits (reflected by some of the Piura participants) are also posed with physical and transportation barriers. If feasible, a door-to-door vaccine campaign should be rolled-out, but it is important to ensure pre-existing trusted sources are used. Community members are used to government supported vector control visiting their houses, so there is potential to pilot adding a vaccination element to existing spraying campaign schedules. Data from Colombia and Singapore found participation in vector control activities was positively associated with dengue vaccine acceptance [62]. Other context factors included differences related to individual characteristics, such as education level and number of people in the household. Individual personalities also play a role, as participants who have participated in past public health programs, such as vector control and COVID-19 vaccine campaigns, are more likely to get a new vaccine.

Complacency in the form of self-perceived risk of dengue influenced participant interest in a dengue vaccine, and dengue naïve individuals or those with mild disease reported a lower self-perception of risk. This is consistent with data from several countries in South America and Southeast Asia that found perceived risk of dengue is positively correlated with vaccine acceptance [62]. It was interesting to note that hearing an unsubstantiated rumor of an adverse COVID-19 outcome was enough to convince someone not to get the vaccine, but personally knowing someone who had dengue didn’t have as strong of an effect for dengue vaccine uptake. There are also implications of hesitation towards larger health goals, as individuals were hesitant towards both the COVID-19 and dengue vaccines, which should be addressed as part of a larger health issue. Dengue education should be revisited, and educational materials should focus on dengue clinical disease, including who is at risk and why. It is well established that second infection with a DENV serotype different from the first infection can progress to a more severe presentation of disease due to antibody-dependent enhancement [64–66], and this is an important message to leverage for individuals who had a mild self-limiting disease to increase their perception of risk.

On this note, an important finding when considering vaccine roll-out is that FGD participants wanted more information on how the vaccine would influence vector control. Vaccine rollouts are often slow and limited supplies of vaccines can lead to some groups getting vaccinated before others. The vaccine is designed to be used in tandem with vector control strategies, so messaging around this needs to be very clear so vaccinated individuals don’t disregard MINSA fumigation and larvicide campaigns, especially since it’s likely not everyone in their neighborhood will be vaccinated at once.

Various key factors emerged from this study that can provide guidance for future vaccine roll out strategies, including the role of misinformation that could affect vaccine hesitancy and strategies to counteract these influences and improve public understanding of vaccine safety and efficacy; the influence of education and socioeconomic level (particularly the finding that higher education, typically associated with better health literacy, was linked to greater hesitancy); the importance of trust in health professionals and local leaders in influencing vaccine acceptance; the need for clear, consistent, and culturally appropriate communication strategies, highlighting the importance of targeted messaging; and finally logistical and physical barriers that could impede access to vaccine uptake.

### Limitations

When this study began, there were no dengue vaccines being used in either study site. There are several factors that impact the generalizability and interpretation of the findings. First, this study took place in specific urban and peri-urban areas of two dengue endemic sites, which may not reflect views across other geographical and socio-economic settings in Peru or Latin America. Second, this study took place two years post the start of the pandemic: attitudes and perceptions might have shifted since then due to evolving health messages or experiences. Third, self-reported data on vaccine acceptance and hesitancy may be subject to social desirability bias; this may be exacerbated by discussing a dengue vaccine which is not yet available. In other words, the study’s focus on attitudes toward a hypothetical dengue vaccine may not capture actual behaviors and decision-making processes once a vaccine becomes available.

## CONCLUSION

A future dengue vaccine will provide much needed protection against a disease that is currently relying on one method of prevention: vector control. The study findings highlight several factors influencing acceptance or refusal of a new dengue vaccine in Peru. Positive attitudes towards a dengue vaccine were associated with experiences of severe dengue, trust in healthcare professionals, and convenience of access, such as house-by-house administration. On the other hand, higher education levels, negative views on the COVID-19 vaccine, and lack of knowledge about dengue transmission, are associated with increased vaccine hesitancy. While most participants expressed interest in the vaccine, concerns about side effects and misinformation, particularly from religious sources, remain significant barriers. Effective communication strategies, tailored outreach, and community engagement, especially in challenging areas like peri-urban regions, are essential for improving vaccine uptake.

## Data Availability

The data analyzed in this study cannot be made publicly available due to ethical and privacy concerns. Focus group transcripts were deidentified, although they still contain information that could potentially be used to identify participants. While the full dataset is not publicly available, researchers with a justified interest in reviewing the data may submit a request to the Tulane University Institutional Review Board (IRB): Tulane University IRB Phone: (504) 988-2665, Fax: (504) 988-4766.

## ACKNOWLEDGEMENTS

The researchers would like to thank all the participants in this study as well as the research team members who assisted with transcribing the audio. THANK SURVEY COLLECTORS

## Financial Disclosure Statement

VAPS supported in part by a research grant from Investigator-Initiated Studies Program of Merck Sharp & Dohme LLC: Merck Investigator Studies Program (MISP) Clinical Research 60875. The opinions expressed in this paper are those of the authors and do not necessarily represent those of Merck Sharp & Dohme LLC. The funders had no role in study design, data collection and analysis, decision to publish, or preparation of the manuscript.

## Author contributions

The conceptualization and study design were conducted by VAPS, ACM, and LM. Data collection instrument design was conducted by JLCC, VAPS, ACM, and LM. Focus groups were conducted by JLCC, ASV, CC, EJRL, JJCL, and CH. Survey data collection was managed by JLCC, ASV, and CC and quality control performed by JJCL and JLCC. Data analysis was performed by JLCC, ACM, and EBO. The manuscript was drafted by EBO, JLCC, VAPS, and ACM. All authors reviewed and approved the final draft of the manuscript.

## Notes

### Competing Interest Statement

The authors have declared no competing interest.

### Author Declarations

Our study protocol was approved by the Institutional Review Boards (IRBs) of PRISMA (CE0882.21) providing local Peruvian approval and the Tulane School of Public Health and Tropical Medicine (2021-1749). The University of California, Davis IRB provided an exemption.

